# Evidence that extracellular HSPB1 contributes to inflammation in alcohol-associated hepatitis

**DOI:** 10.1101/2024.09.06.24313193

**Authors:** Anne-Marie C Overstreet, McKenzie Burge, Annette Bellar, Megan McMullen, Douglas Czarnecki, Emily Huang, Vai Pathak, Chelsea Finney, Raveena Vij, Srinivasan Dasarathy, Jaividhya Dasarathy, David Streem, Nicole Welch, Daniel Rotroff, Adam M Schmitt, Laura E Nagy, Jeannette S Messer

**Author notes:** **Corresponding author:** Jeannette S Messer.

## Abstract

**Background and aims:** Alcohol-associated hepatitis (AH) is the most life-threatening form of alcohol-associated liver disease (ALD). AH is characterized by severe inflammation attributed to increased levels of ethanol, microbes or microbial components, and damage-associated molecular pattern (DAMP) molecules in the liver. HSPB1 (Heat Shock Protein Family B (Small) Member 1; also known as Hsp25/27) is a DAMP that is rapidly increased in and released from cells experiencing stress, including hepatocytes. The goal of this study was to define the role of HSPB1 in AH pathophysiology.

**Methods:** Serum HSPB1 was measured in a retrospective study of 184 heathy controls (HC), heavy alcohol consumers (HA), patients with alcohol-associated cirrhosis (AC), and patients with AH recruited from major hospital centers. HSPB1 was also retrospectively evaluated in liver tissue from 10 HC and AH patients and an existing liver RNA-seq dataset. Finally, HSPB1 was investigated in a murine Lieber-DeCarli diet model of early ALD as well as cellular models of ethanol stress in hepatocytes and hepatocyte-macrophage communication during ethanol stress.

**Results:** Circulating HSPB1 was significantly increased in AH patients and levels positively correlated with disease-severity scores. Likewise, HSPB1 was increased in the liver of patients with severe AH and in the liver of ethanol-fed mice. *In vitro*, ethanol-stressed hepatocytes released HSPB1, which then triggered TNFα-mediated inflammation in macrophages. Anti-HSPB1 antibody prevented TNFα release from macrophages exposed to media conditioned by ethanol-stressed hepatocytes.

**Conclusions:** Our findings support investigation of HSPB1 as both a biomarker and therapeutic target in ALD. Furthermore, this work demonstrates that anti-HSPB1 antibody is a rational approach to targeting HSPB1 with the potential to block inflammation and protect hepatocytes, without inactivating host defense.

**GRAPHICAL ABSTRACT:** 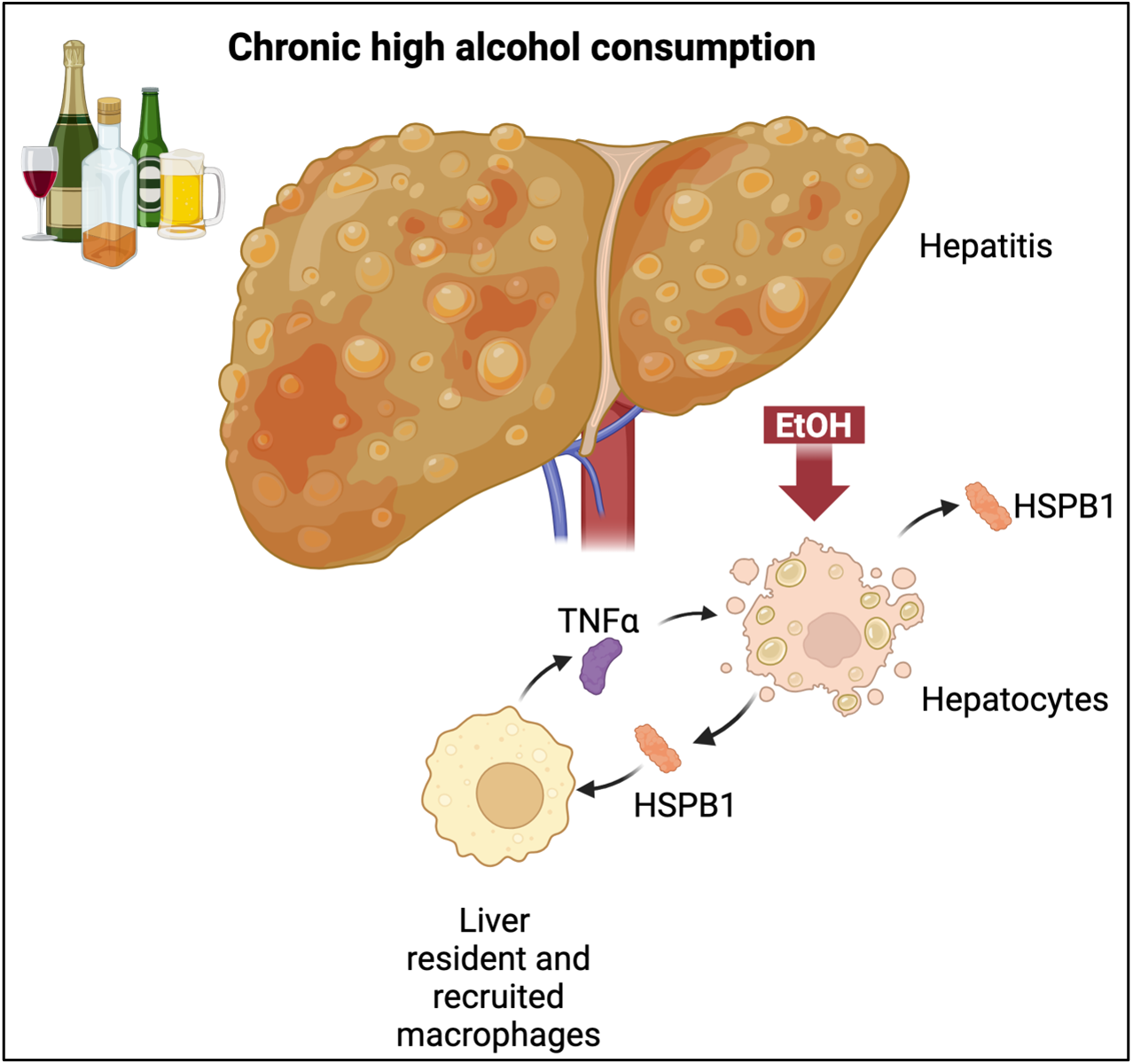

**HIGHLIGHTS:**

- HSPB1 is significantly increased in serum and liver of patients with alcohol-associated hepatitis.
- Ethanol consumption leads to early increases in HSPB1 in the mouse liver.
- Hepatocytes subjected to ethanol stress release HSPB1 into the extracellular environment where it activates TNFα-mediated inflammation in macrophages.
- Anti-HSPB1 antibody blocks hepatocyte-triggered TNFα in a model of hepatocyte-macrophage communication during ethanol stress.

## INTRODUCTION

Alcohol-associated liver disease (ALD) has reached a global prevalence of 4.8% and is now the most common cause of death due to excessive alcohol consumption in the United States (US).^1, 2^ In the US, ALD mortality increased 23% between 2019 and 2020, and claimed an estimated 45,290 American lives in 2022.^1, 3^ ALD has also become the leading indication for liver transplantation in the US, with a 63% increase in the number of patients listed for transplant between 2007 and 2017.^4, 5^ The term ALD broadly refers to any type of alcohol-induced liver damage and encompasses steatosis, steatohepatitis, cirrhosis, and hepatocellular carcinoma.^6^ In general, higher levels of alcohol consumption lead to more severe disease, but transitions between types of disease are unpredictable and ALD may go unrecognized until it becomes severe. These factors make it imperative to understand the pathophysiology of ALD in order to develop reliable diagnostics that predict risk of progression and therapies able to prevent severe forms of disease.

Alcohol-associated hepatitis (AH) is the most severe and life-threatening manifestation of ALD.^7^ Mortality in severe AH approaches 50% within three months of diagnosis and rises to 70% in patients that do not respond to corticosteroids.^8^ Patients with AH present with rapid onset signs of liver failure and systemic inflammation.^9^ Liver biopsy in these patients typically reveals steatosis, hepatocyte ballooning, Mallory-Denk hyaline inclusions, fibrosis, and infiltration by innate and adaptive immune cells.^9^ The inflammatory cell infiltrate is accompanied by high levels of pro-inflammatory cytokines such as TNFα (tumor necrosis factor alpha), IL-1β (interleukin-1 beta), and IL-6 (interleukin-6).^9^ These factors suggest that AH is an acute inflammatory disease superimposed on chronic ALD.^6^ They also argue that targeting inflammation could slow or reverse liver damage and decrease AH-associated mortality.

Inflammation results from both direct and indirect effects of alcohol on the liver. In the normal liver, tissue resident macrophages, called Kupffer cells, line the sinusoids and react to microbes, microbial components, and microbial products transmitted to the liver from the gastrointestinal (GI) tract.^10^ These microbe-associated molecular pattern molecules (MAMPs) are detected by innate immune receptors on the surface or in the cytosol of macrophages and activate signaling cascades that lead to production and release of cytokines and chemokines. In the absence of alcohol, Kupffer cells are exposed to low levels of MAMPs, and inflammation is limited in both extent and duration.^11^ In contrast, alcohol can directly activate production and release of AH-associated cytokines, such as TNFα, from Kupffer cells or macrophages recruited into the liver.^12^ Alcohol also damages the gut mucosal barrier, allowing increased translocation of MAMPs to the liver where they activate production of the same suite of pro-inflammatory cytokines and chemokines in Kupffer cells or recruited macrophages.^13^ Finally, ethanol stressed hepatocytes can contribute to macrophage activation through release of damage-associated molecular pattern molecules (DAMPs). DAMPs are intracellular proteins that are naturally found at high levels inside cells or that are rapidly upregulated in response to stress. These danger signals are actively secreted from ethanol stressed cells or released into the extracellular environment secondary to cell death.^14^ In the extracellular environment, DAMPs activate pattern recognition receptors (PRR) on macrophages, which leads to production of the same cytokines and chemokines that macrophages produce in response to MAMPs.^15^

One DAMP of particular interest in AH inflammation is HSPB1 (Heat Shock Protein Family B (Small) Member 1; also known as Hsp25/27). HSPB1 protein levels are rapidly increased in a wide range of cells subjected to MAMPs or oxidative stress.^16–18^ In these cells, HSPB1 directly and indirectly counteracts cell damage and programmed cell death mechanisms, allowing them to survive in otherwise deadly conditions.^19, 20^ Although HSPB1 has not been systematically evaluated in ALD, it has been implicated in alcohol-associated disease in other ways. Hepatocytes are primarily responsible for ethanol metabolism in the liver and this metabolism generates reactive oxygen species that accumulate and cause oxidative stress and cell damage with high or prolonged alcohol use.^21^ There is a large body of literature demonstrating that HSPB1 acts to prevent programmed cell death in cells experiencing oxidative stress.^22^ A role for HSPB1 in cellular responses to ethanol is further supported by studies in astrocytes, which showed that ethanol challenge leads to increased HSPB1 gene expression in in these cells.^23^ Extracellular HSPB1 is of particular interest in ALD since patients with hepatocellular carcinoma have increased levels of HSPB1 and anti-HSPB1 antibodies in their blood; increases that were linked to a history of alcohol use in some studies.^24–28^ HSPB1 is reportedly released from stressed cells, including hepatocytes to activate PRR on adjacent cells.^29–32^ HSPB1-mediated PRR activation then leads to production and release of the AH-associated cytokines TNFα and IL-1β from macrophages.^32, 33^

These factors led us to examine HSPB1 in AH, with the goal of understanding the role that this protein plays in inflammation and liver damage caused by alcohol consumption. We found that patients with AH have significantly higher levels of HSPB1 in both their blood and liver than liver disease free controls or patients with alcohol-associated cirrhosis (AC). This suggested that increases in HSPB1 were related to the higher level of inflammation in AH. In a murine model of chronic alcohol consumption (CAC), HSPB1 was increased in the liver prior to significant liver damage, meaning that it does not result from inflammation. Similar to our human and animal studies, HSPB1 protein was rapidly increased in and released from hepatocytes stressed with ethanol *in vitro*. In the extracellular environment, HSPB1 promoted inflammation by triggering TNFα release from macrophages. The importance of HSPB1 in ethanol-activated inflammation was reinforced by the fact that anti-HSPB1 antibody almost completely mitigated TNFα release from macrophages treated with media conditioned by ethanol stressed hepatocytes. Together, these findings establish a key role for HSPB1 in the inflammation of AH and identify it as a potential biomarker and therapeutic target for this intractable disease.

## RESULTS

### Circulating HSPB1 is increased in ALD patients with the highest levels in severe AH

Predicting the course of ALD in a given patient is extremely challenging. Nearly all patients consuming high levels of alcohol develop steatosis and many never progress to more severe disease.^9^ However, patients can progress to life-threatening disease quickly and without any prior indication. This makes development of biomarkers that can identify patients at risk of progression vital in ALD. The ideal biomarker would be amenable to non-invasive monitoring, differentiate liver disease caused by alcohol use from other causes, predict risk of acute destabilization, and be targetable to prevent severe disease.

Identification of HSPB1 as a potential DAMP and biomarker of inflammation in AH led us to retrospectively measure HSPB1 in serum from healthy controls (HC), heavy alcohol consumers (HA), patients with AC, and patients with AH (**Table 1**). Levels of HSPB1 were similar in the groups without liver disease (HC and HA). However, HSPB1 was significantly higher in patients with ALD (AC and AH) versus those without liver disease (**Fig. 1A**). Comparison of serum HSPB1 concentrations between AC and AH further revealed that HSPB1 was significantly higher in AH than AC. We next investigated relationships between disease severity and HSPB1 in the ALD groups (**Table 2**). There was no correlation between HSPB1 and disease severity in AC and no difference between HSPB1 in patients with severe versus mild-moderate AC (**Fig. 1B, C**). However, within the AH group, HSPB1 increased as disease severity increased and levels of HSPB1 were significantly higher in severe versus mild-moderate disease (**Fig. 1D, E**). HSPB1 levels were not related to age or sex, despite literature describing HSPB1 as an estrogen responsive gene (**Fig. 1F, G**).^31^ Thus, high circulating HSPB1 is a feature of severe AH.

**Figure 1.**
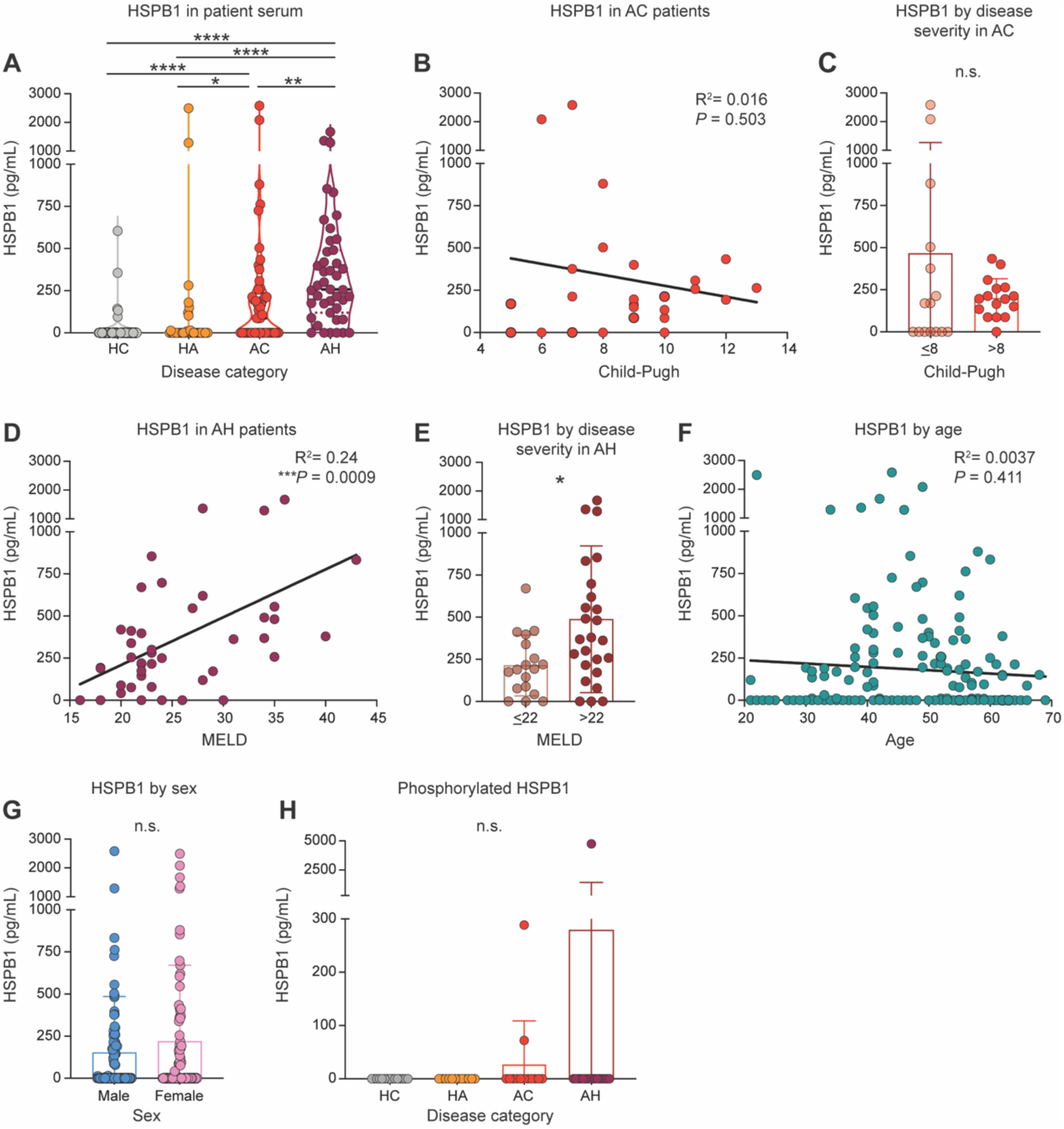
Circulating HSPB1 is increased in ALD patients. **A**, Violin plot of HSPB1 concentration in patient sera measured by ELISA. **B**, Linear regression of HSPB1 concentration and Child-Pugh score of AC patients. **C**, HSPB1 concentration in AC patients with mild-moderate (≤8) vs. severe (>8) disease scores (Child-Pugh). **D**, Linear regression of HSPB1 concentration vs. MELD score of AH patients. **E**, HSPB1 concentration in AH patients with mild-moderate (≤22) vs. severe (>22) disease scores (MELD). **F**, Linear regression of HSPB1 concentration vs. age in all patients. **G**, HSPB1 concentration in male vs. female patients. **H**, Phosphorylated HSPB1 concentration in patient sera measured by ELISA. All data plotted as mean ± SEM. A,H. Kruskal-Wallace test with a Dunn’s multiple comparisons post-hoc test. B, D, F. Simple Linear Regression. C, E, G. Mann-Whitney test. HC, healthy control; HA, high alcohol consumer; AC, alcohol-associated cirrhosis; AH, alcohol-associated hepatitis. n.s. *P* > 0.05, * *P* ≤ 0.05, ** *P* ≤ 0.01, *** *P* ≤ 0.001, **** *P* ≤ 0.0001.

**Table 1.**
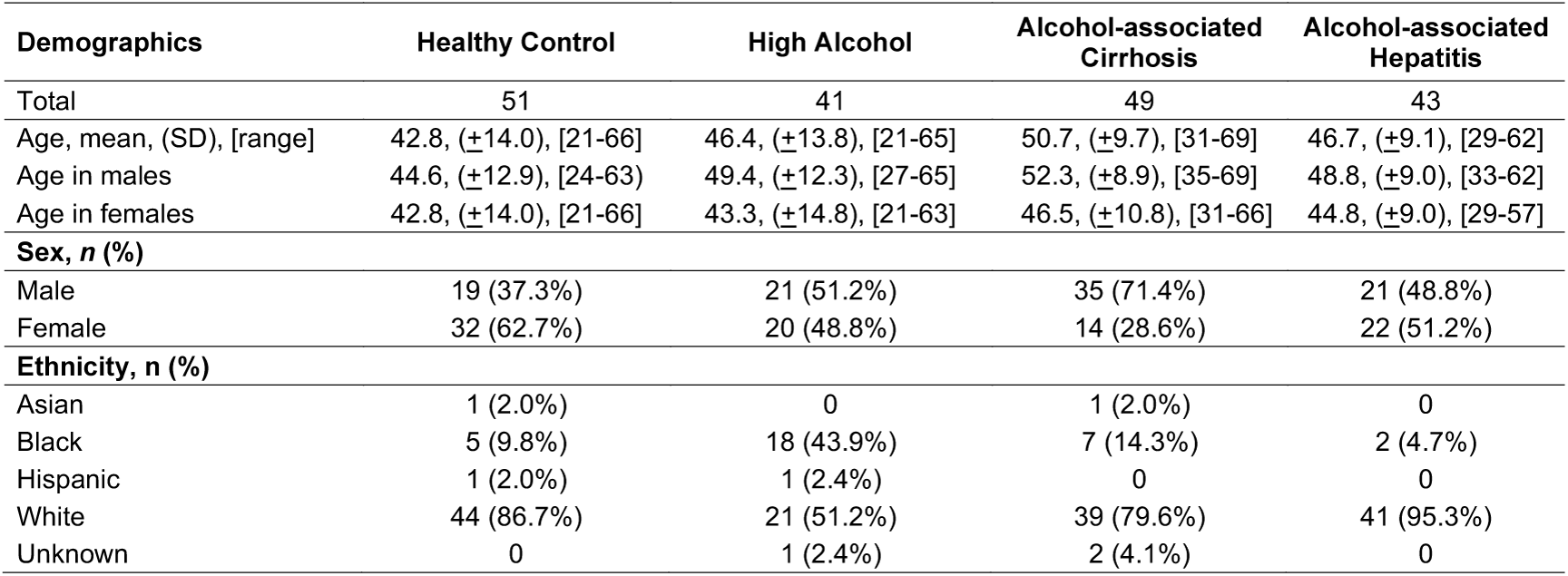
Patient demographics in studies of circulating HSPB1.

**Table 2.** Patient demographics and clinical data in studies of circulating HSPB1 and disease severity.

| Demographics | Alcohol-associated Cirrhosis | Alcohol-associated Hepatitis |
| --- | --- | --- |
| Total | 31 | 43 |
| Age, mean, (SD), [range] | 50.6, ($\pm 9.9$), [31-68] | 46.7, ($\pm 9.1$), [29-62] |
| Age in males | 52.2, ($\pm 9.0$), [38-68] | 48.8, ($\pm 9.0$), [33-62] |
| Age in females | 47.8, ($\pm 11.1$), [31-66] | 44.8, ($\pm 9.0$), [29-57] |
| Score | Child-Pugh | MELD |
| Score, mean, (SD), [range] | 8.4, ($\pm 2.2$), [5-13] | 25.8, ($\pm 6.5$), [16-43] |
| Score in males | 8.5, ($\pm 2.3$), [5-13] | 27.5, ($\pm 7.3$), [18-43] |
| Score in females | 8.4, ($\pm 2.2$), [5-12] | 24.1, ($\pm 5.2$), [16-36] |
| Sex, n (%) |  |  |
| Male | 20 (64.5%) | 21 (48.8%) |
| Female | 11 (35.5%) | 22 (51.2%) |
| Ethnicity, n (%) |  |  |
| Asian | 0 | 0 |
| Black | 1 (3.2%) | 2 (4.7%) |
| Hispanic | 0 | 0 |
| White | 30 (96.8%) | 41 (95.3%) |
| Unknown | 0 | 0 |

Several post-translational modifications have been described for intracellular HSPB1, including phosphorylation. Human HSPB1 contains three serines capable of accepting a phospho-group, serine 15, serine 78, and serine 82.^34^ Phosphorylation of these serines determines oligomerization and distribution of HSPB1 inside cells, but the role of phosphorylation in extracellular functions is far less clear. Serine 15 phosphorylation is associated with HSPB1 distribution to insoluble cell components during hydrogen peroxide induced oxidative stress, making it unlikely to be found in extracellular HSPB1 during ethanol stress.^18^ This made us focus on HSPB1 phosphorylation at serines 78 and 82 in the patient blood (**Table 3**). Of the tested samples, only 3 of 57 were positive for phosphorylated-HSPB1, two of the positive samples were in the AC group and one was in the AH group (**Fig. 1H**). All of the samples positive for phosphorylated HSPB1 were also positive for total HSPB1. This suggests that extracellular HSPB1 is only rarely phosphorylated in ALD patients and so, phosphorylation is not likely to be involved in its extracellular functions in AH.

**Table 3.** Patient demographics and clinical data in studies of phosphorylated HSPB1 in serum.

| Demographics | Healthy Control | High Alcohol | Alcohol-associated Cirrhosis | Alcohol-associated Hepatitis |
| --- | --- | --- | --- | --- |
| Total | 15 | 12 | 13 | 17 |
| Age, mean, (SD), [range] | 40.4, ($\pm 12.7$), [23-62] | 42.3, ($\pm 12.8$), [24-63] | 47.9, ($\pm 8.0$), [35-60] | 45.6, ($\pm 9.4$), [30-60] |
| Age in males | 43.0, ($\pm 12.4$), [25-62] | 47.0, ($\pm 12.8$), [30-63] | 49.3, ($\pm 5.4$), [41-56] | 45.9, ($\pm 9.6$), [33-60] |
| Age in females | 37.4, ($\pm 13.2$), [23-62] | 32.8, ($\pm 6.0$), [24-37] | 46.6, ($\pm 10.0$), [35-60] | 44.6, ($\pm 9.6$), [30-55] |
| Score |  |  | Child-Pugh | MELD |
| Score, mean, (SD), [range] | | | 8.4, ($\pm 3.4$), [5-13] | 22.3, ($\pm 3.9$), [16-30] |
| Score in males | | | 8.7, ($\pm 2.4$), [7-13] | 23.4, ($\pm 4.2$), [18-30] |
| Score in females | | | 8.1, ($\pm 2.5$), [5-12] | 21.1, ($\pm 3.2$), [16-27] |
| Sex, n (%) |  |  |  |  |
| Male | 8 (53.3%) | 8 (66.7%) | 6 (46.2%) | 8 (47.1%) |
| Female | 7 (46.7%) | 4 (33.3%) | 7 (53.8%) | 9 (52.9%) |
| Ethnicity, n (%) |  |  |  |  |
| Asian | 0 | 0 | 0 | 0 |
| Black | 2 (13.3%) | 2 (16.7%) | 0 | 0 |
| Hispanic | 1 (6.7%) | 0 | 0 | 0 |
| White | 12 (80.0%) | 9 (75.0%) | 13 (100%) | 17 (100%) |
| Unknown | 0 | 1 (8.3%) | 0 | 0 |

### HSPB1 is increased in the liver of patients with AH

We next considered the source of circulating HSPB1 in patients with severe AH. The liver is responsible for the majority of ethanol metabolism and is subject to high levels of oxidative stress generated by the highly reactive byproducts of this metabolism.^35^ A large body of literature has demonstrated that HSPB1 is cytoprotective in cells experiencing oxidative stress.^36^ HSPB1 release from stressed hepatocytes has also been reported.^29^ This suggested that the liver was the most likely source of circulating HSPB1 in the AH patients.

In the United States, very few ALD patients undergo diagnostic liver biopsy, so large numbers of liver samples from these patients are difficult to obtain. We were fortunate to acquire banked liver lysates from five patients with severe AH and five healthy controls (**Table 4**). These samples were obtained during liver transplantation from patients with severe AH and healthy donor livers. There was a clear difference in HSPB1 between HC and AH, with HSPB1 significantly higher in AH than HC (**Fig. 2A, B**). We also utilized existing RNA-seq datasets to evaluate HSPB1 gene expression in the livers of patients with AH of varying severity or other types of liver disease. In non-severe and severe AH (liver biopsies or explanted liver), HSPB1 gene expression was significantly higher than both HC and any other type of liver disease (**Fig. 2C, D, E**). Interestingly, metabolic dysfunction-associated steatotic liver disease (MASLD) was not associated with increased HSPB1 gene expression. This is similar to reports in the literature that HSPB1 in circulation does not differ between pediatric healthy controls and patients with MASLD and that ballooning degeneration is associated with decreased HSPB1 gene expression and protein in livers from adult MASLD patients.^37, 38^ Taken together, these data identify the liver as an important source of circulating HSPB1 in AH and suggest that high levels of HSPB1 in the liver are linked to ALD, but not MASLD.

**Figure 2.**
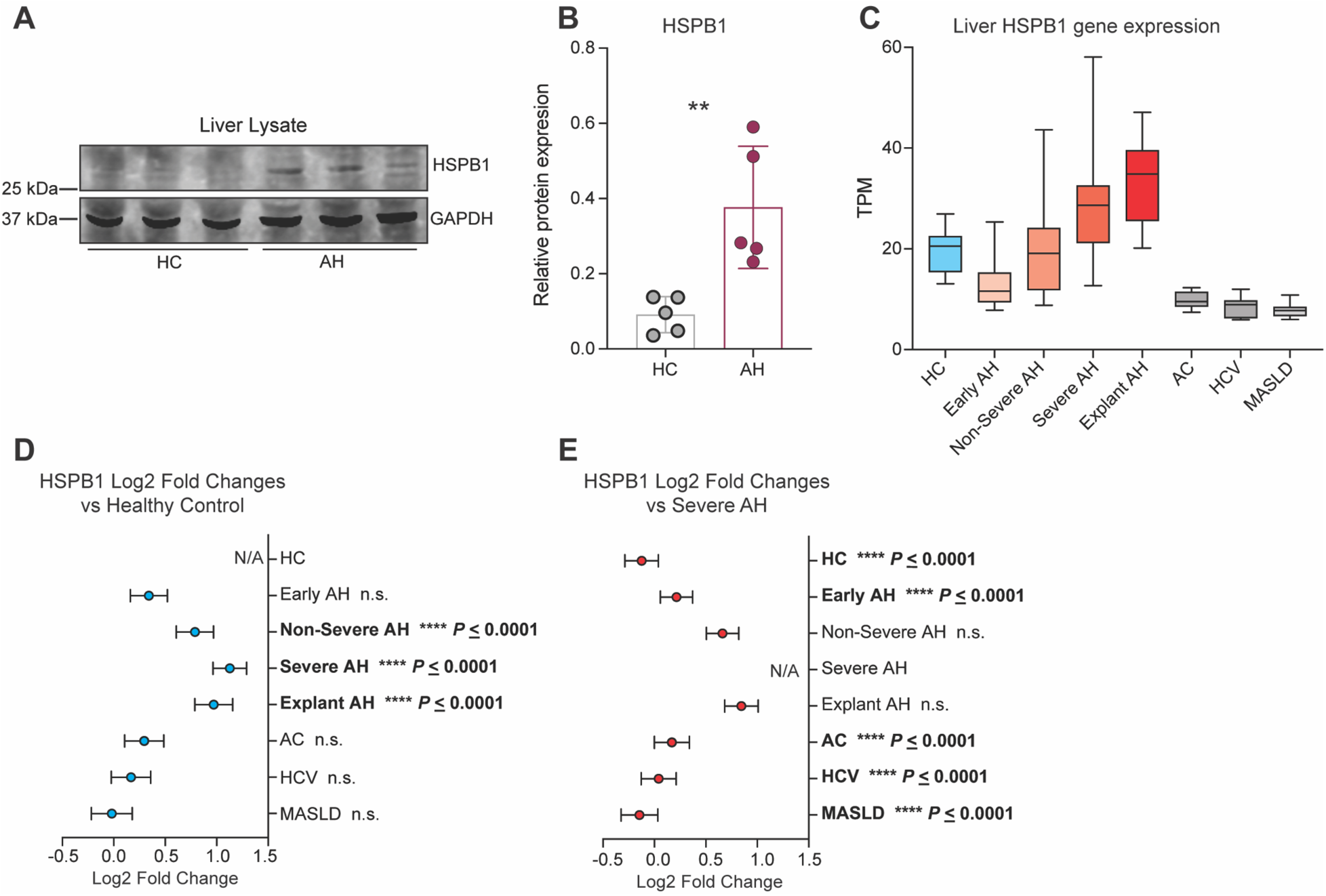
HSPB1 is increased in the liver of patients with AH. **A**, Representative immunoblot for HSPB1 and GAPDH in HC and AH liver samples. **B**, HSPB1 protein concentration relative to GAPDH as measured by densitometry in immunoblots represented in (**A**). **C**, HSPB1 gene expression in RNA-seq analysis of liver from HC, AH, AC, MASLD, and HCV. **D**, Forrest plot depicting Log2 fold changes in HSPB1 expression in samples described in (**C**). Patients with various types of liver disease were compared to healthy controls. **E**, Forrest plot depicting Log2 fold changes in HSPB1 expression in samples described in (**C**). Patients with various types of liver disease were compared to patients with severe AH. Data plotted as mean ± SD, except C,D,E which is mean ± SEM. B. Mann-Whitney test. C, D. DeSeq2. HC, healthy control; AH alcohol-associated hepatitis, MASLD, metabolic dysfunction associated with steatotic liver disease; HCV, hepatitis C virus; TPM, transcripts per million. n.s. *P* > 0.05, * *P* ≤ 0.05, ** *P* ≤ 0.01, *** *P* ≤ 0.001, **** *P* ≤ 0.0001.

**Table 4.** Patient demographics and clinical data for samples used for HSPB1 liver lysate immunoblot.

| Demographics | Healthy Control | Alcohol-associated Hepatitis |
| --- | --- | --- |
| Total | 5 | 5 |
| Age, mean, (SD) | 45.6, (±6.7) | 40.8, (±3.5) |
| **Sex, *n* (%)** |  |  |
| Male | 3 (60%) | 3 (60%) |
| Female | 2 (40%) | 2 (40%) |
| **Clinical Data** |  |  |
| Decompensation (*n*, (%)) | N/A | 5 (100%) |
| Sepsis (*n*, (%)) | N/A | 0 (0%) |
| Maddrey's discriminant function | N/A | 102.5 (±27.7) |
| Total bilirubin (mg/dL) | N/A | 25.2 (±5.1) |
| Prothrombin time (sec) | N/A | 32.4 (±5.7) |

### Chronic ethanol consumption increases HSPB1 in murine liver

HSPB1 could be increased in the liver secondary to damage, or it could be contributing to damage. To distinguish between these possibilities, we examined livers using a murine model of CAC. Mice were fed the Lieber-DeCarli liquid diet with increasing amounts of ethanol over 25 days or a calorie-matched diet supplemented with maltodextrin (**Fig. 3A**).^39^ The final concentration of ethanol in this model is 6% and that concentration is maintained for one week. Controls were pair-fed, meaning that their calorie intake was adjusted daily to match that of a paired ethanol-consuming mouse. This model replicates early or mild ALD. We chose this model because our goal was to determine whether increased liver HSPB1 results from liver damage or precedes and therefore potentially causes liver pathology seen in AH.

**Figure 3.**
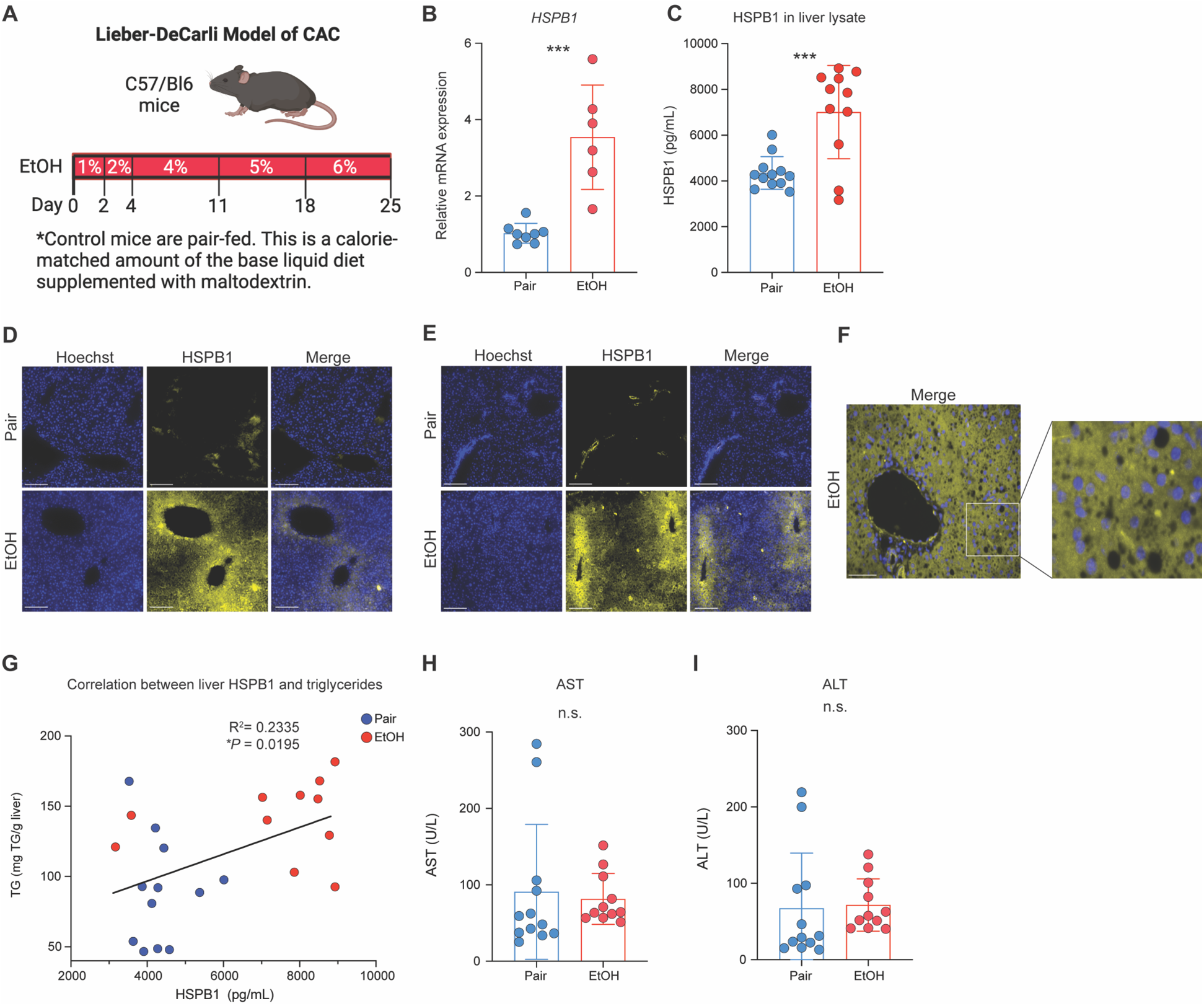
Chronic ethanol consumption increases HSPB1 in murine liver. **A**, Illustration of murine model of CAC. **B**, Relative gene expression of HSPB1 in the liver of Pair and EtOH mice measured by qRT-PCR. **C**, HSPB1 concentration in the liver of Pair and EtOH mice measured by ELISA. **D**&**E**, Immunostaining for HSPB1 (yellow) and DNA (Hoechst, blue) in pair- and EtOH-fed mice. Scale bar = 100 μm. Images in (**D**) are focused on periportal hepatocytes and images in (**E**) are focused on pericentral hepatocytes. Images are representative of 12 Pair and 11 EtOH mice. **F**, High-magnification image of HSPB1 (yellow) and DNA (Hoechst, blue) immunofluorescence in the liver of EtOH mice. Scale bar = 50 μm. **G**, Linear regression analysis of liver HSPB1 and triglyceride levels in Pair and EtOH mice. **H**, AST levels in serum from Pair and EtOH mice. **I**, ALT levels in serum from Pair and EtOH mice. Data are plotted as mean ± SD. B, C, H, I. Unpaired T-test. G. Simple Linear Regression. CAC, chronic alcohol consumption; Pair, pair-fed mice; EtOH, ethanol-fed mice; TG, triglycerides; AST, aspartate aminotransferase; ALT, alanine aminotransferase. n.s. *P* > 0.05, * *P* ≤ 0.05, ** *P* ≤ 0.01, *** *P* ≤ 0.001, **** *P* ≤ 0.0001.

Ethanol-fed (EtOH) mice had significantly higher levels of both HSPB1 mRNA and protein in their livers than pair-fed (Pair) controls (**Fig. 3B, C**). Immunofluorescence staining of liver sections from these mice identified weak HSPB1 staining in periportal regions and high HSPB1 staining around central veins in EtOH mice (**Fig. 3D, E**). Periportal hepatocytes are exposed to high levels of gut-derived components, including MAMPs.^40^ Ethanol consumption, especially at high levels, damages the gut epithelium, allowing increased translocation of microbes, microbial components, or microbial products across the barrier and into the liver through portal circulation.^41^ HSPB1 production in response to MAMPs such as lipopolysaccharide and flagellin is well-documented and is likely responsible for the weak periportal HSPB1 staining in the EtOH mice.^16, 17^ In contrast, HSPB1 staining was pronounced in pericentral hepatocytes, which are primarily responsible for ethanol metabolism in the liver.^42^ Of note, hepatocytes positive for HSPB1 were also filled with lipid droplets and HSPB1 levels in the liver were positively correlated with triglyceride levels in the blood of these mice (**Fig. 3F, G**). This is in keeping with a report that HSPB1 knockout mice have lower levels of triglycerides in their blood.^43^ The presence of HSPB1 in the ethanol-stressed liver prior to damage was reinforced by the fact that circulating aspartate aminotransferase (AST) and alanine aminotransferase (ALT) were similar in EtOH and Pair mice (**Fig. 3H, I**). Since HSPB1 is increased in the ethanol-stressed liver prior to the onset of significant damage, the increased HSPB1 is more likely to be a cause of damage than the result of damage.

### Ethanol stressed hepatocytes release HSPB1 into the extracellular environment

Findings from patients and mice chronically consuming alcohol identified the liver, and specifically hepatocytes, as the likely source of circulating HSPB1 in AH. To verify ethanol activated HSPB1 release from hepatocytes, we performed *in vitro* studies of ethanol challenge using hepatocyte cell lines.

Ethanol exposure rapidly and robustly increased HSPB1 protein in HepG2 cells, even before HSPB1 mRNA levels increased (**Fig. 4A, B, C**). This type of rapid upregulation without the need for increased transcription has been reported for heat shock proteins, including HSPB1 with other types of cell stress.^44, 45^ It also fits with the extensively documented roles that HSPB1 plays in cellular survival during many different types of stress, including oxidative stress. Since our primary interest in hepatocyte produced HSPB1 was in HSPB1 release from hepatocytes, we investigated whether ethanol challenge led to increased amounts of HSPB1 in the hepatocyte media. Exposure to ethanol led to an approximately 3-fold increase in extracellular HSPB1 within 1 hr. (**Fig. 4D**). This increase was triggered in response to ethanol metabolism, not cell membrane damage, since inhibiting ethanol metabolism with 4-methylpyrazole (4-MP) completely eliminated HSPB1 release in response to ethanol challenge (**Fig. 4E, F**). We performed these experiments in two hepatocyte cell lines, HepG2 (human) and AML12 (murine), to account for any cell-line related effects on ethanol metabolism or HSPB1 production and release. Thus, ethanol metabolism leads to rapid increases in both intracellular HSPB1 and HSPB1 released from hepatocytes.

**Figure 4.**
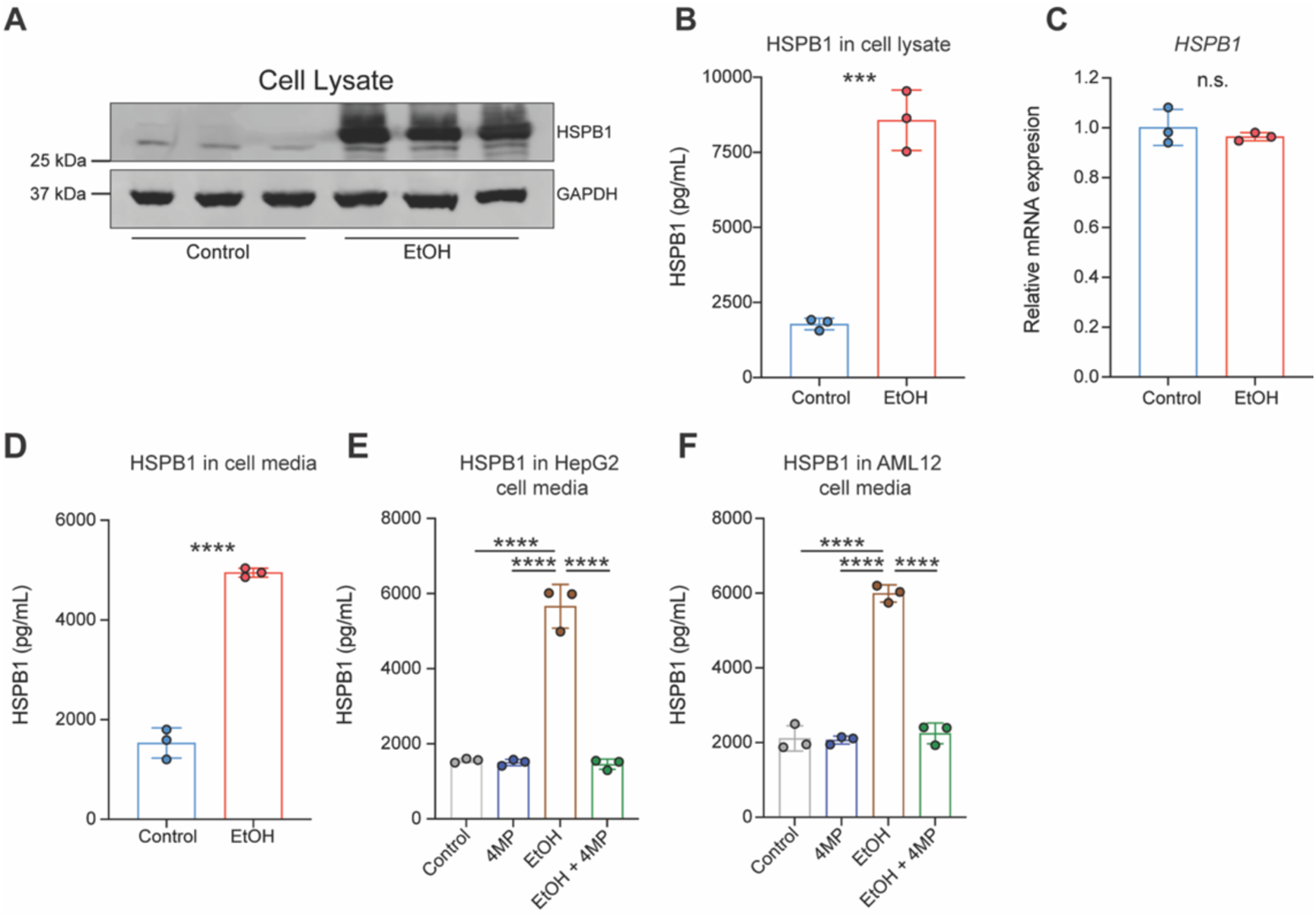
HSPB1 is increased in EtOH-stressed hepatocytes and released into the extracellular environment. **A**, Immunoblotting for HSBP1 and GAPDH in HepG2 cells exposed to vehicle control or 2.5 mM (1.5%) EtOH for 1 hr. **B**, Intracellular HSPB1 concentration measured in cell lysates of HepG2 cells exposed to vehicle control or 2.5 mM EtOH for 1 hr. by ELISA. **C**, HSPB1 gene expression relative to GAPDH in HepG2 cells exposed to vehicle control or 2.5 mM EtOH for 1 hr. by qRT-PCR. **D**, Extracellular HSPB1 concentration measured in culture media from HepG2 cells exposed to vehicle control or 2.5 mM EtOH for 1 hr. by ELISA. **E**, Extracellular HSPB1 concentration measured in cell media from HepG2 cells exposed to vehicle control, 5 mM 4MP, 2.5 mM EtOH, or 2.5 mM EtOH and 5 mM 4MP for 1 hr. by ELISA. **F**, Extracellular HSPB1 concentration measured in cell media from AML12 cells exposed to vehicle control, 5 mM 4MP, 2.5 mM EtOH, or 2.5 mM EtOH and 5 mM 4MP for 1 hr. by ELISA. Data are plotted as mean ± SD. B, C, D. Unpaired T-test. E, F. One-way ANOVA with a Tukey’s multiple comparisons post-hoc test. n.s. *P* > 0.05, * *P* ≤ 0.05, ** *P* ≤ 0.01, *** *P* ≤ 0.001, **** *P* ≤ 0.0001.

### HSPB1 released from ethanol stressed hepatocytes promotes liver-damaging inflammation

In the extracellular environment, HSPB1 functions as a DAMP to amplify macrophage inflammatory responses, including those involving TNFα.^32, 33^ TNFα is of particular interest since it has been implicated in liver damage and hepatocyte death in patients with AH.^46^ This cytokine is also reportedly increased in plasma from liver disease patients with elevated HSPB1 in circulating lymphomonocytes.^47^ Despite minimal changes to the hepatic parenchyma in our EtOH mice receiving the Lieber-DeCarli diet, there was a higher level of TNFα in the liver of these mice than Pair controls (**Fig. 5A**). Furthermore, the level of TNFα was related to the level of HSPB1 in the liver in these mice (**Fig. 5B**).

**Figure 5.**
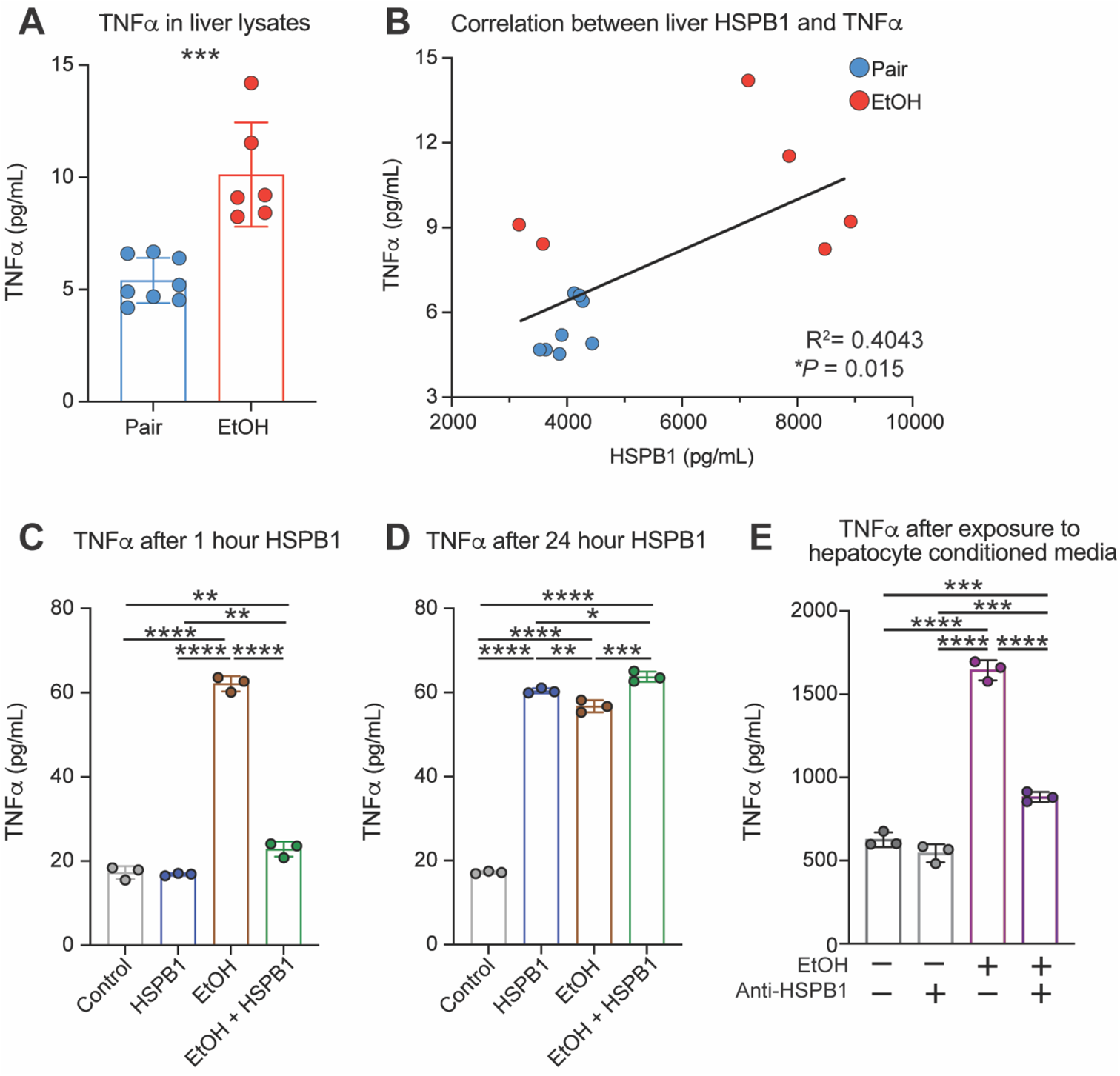
Extracellular HSPB1 promotes liver-damaging inflammation. **A**, TNFα concentration in liver lysates from of Pair and EtOH mice measured by ELISA. **B**, Linear regression analysis of liver HSPB1 and TNFα in Pair and EtOH mice. **C**, TNFα concentration measured in RAW264.7 cells after 1 hr. of treatment with 3 μM HSPB1, 2.5 mM (1.5%) EtOH, or both 2.5 mM EtOH and 3 μM HSPB1 by ELISA. **D**, TNFα concentration measured in RAW264.7 cells after 24 hr. of treatment with 3 μM HSPB1, 2.5 mM EtOH, or both 2.5 mM EtOH and 3 μM HSPB1 by ELISA. **E**, TNFα concentration in RAW264.7 cells after exposure to EtOH stressed HepG2 (2.5 mM EtOH for 24 hr.) conditioned media with or with or without 2.5 mg/ml anti-HSPB1 antibody measured by ELISA. Data are plotted as mean ± SD. A. Unpaired T-test. B. Simple Linear Regression. C, D, E. One-ANOVA with a Tukey’s multiple comparisons post-hoc test. n.s. *P* > 0.05, * *P* ≤ 0.05, ** *P* ≤ 0.01, *** *P* ≤ 0.001, **** *P* ≤ 0.0001.

Macrophages are the primary producers of pro-inflammatory cytokines, including TNFα in the ethanol-stressed liver.^12^ Our finding that ethanol stress triggers HSPB1 release from hepatocytes led us to consider that extracellular HSPB1 could act on nearby macrophages to amplify TNFα secretion during CAC. As previously reported, ethanol robustly increased TNFα secretion by macrophages within one hour of exposure (**Fig. 5C**).^48^ In contrast, TNFα secretion was unchanged in HSPB1 treated macrophages at 1 hour. However, at 24 hours, TNFα secretion was significantly increased in HSPB1 treated macrophages and levels were essentially identical to ethanol-exposed macrophages (**Fig. 5D**). The combination of HSPB1 and ethanol increased TNFα release slightly over each component alone. These data suggest that the mechanisms and kinetics of TNFα secretion elicited by HSPB1 and ethanol differ, but not the inflammatory outcome. They also demonstrate that macrophages need never encounter ethanol or MAMPs directly to activate inflammation during CAC. HSPB1 is sufficient to activate TNFα mediated inflammation and has the potential to amplify inflammation triggered by either ethanol or MAMPs.

We next investigated whether blocking extracellular HSPB1 is sufficient to inhibit TNFα release from macrophages exposed to ethanol stressed hepatocytes. Media conditioned by ethanol stressed HepG2 cells was used to replicate the natural form and amount of HSPB1 released from hepatocytes in these experiments. We added anti-HSPB1 antibody to the hepatocyte conditioned media, then added the treated media to macrophages, and measured TNFα release. The addition of anti-HSPB1 antibody reduced TNFα release from macrophages significantly (**Fig. 5E**). Overall, our data suggest that HSPB1 is an important initiator of TNFα dependent inflammation and hepatocyte death in AH. They also show that anti-HSPB1 antibody is a rational therapeutic strategy to protect the ethanol stressed liver.

## DISCUSSION

Inflammation destabilizes ALD, leading to disease progression and AH. In this study, we discovered that HSPB1 is increased in the liver and blood of patients with severe AH. We also demonstrated that HSPB1 is released from ethanol stressed hepatocytes and triggers inflammation linked with progression to AH and increased mortality in ALD. Extracellular HSPB1 has been identified as a DAMP in published literature. For example, cardiomyocytes subjected to ischemia and reperfusion injury have been shown to release HSPB1 to activate toll like receptor 2/4 on endothelial cells.^49^ Extracellular HSPB1 is also reported to trigger TNFα inflammation in macrophages.^32, 33^ Our work shows that HSPB1 fits the classical profile of a DAMP in ALD, in that it activates macrophages when released from ethanol stressed cells. However, it is unlike many other DAMPs, in that it normally present at low levels and is rapidly increased in response to ethanol. In CAC, this means that it is released from cells that have encountered ethanol or evidence of ethanol toxicity (i.e., MAMPs) and upregulated HSPB1. Release of DAMPs is generally considered to be a product of dead cells.^15^ However, there is ample evidence that HSPB1 is secreted by live cells, including hepatocytes.^29, 50^

This work not only adds HSPB1 to the list of DAMPs identified in ALD, it also demonstrates that this protein is a cause of TNFα mediated inflammation associated with the progression to AH. Inflammation was established as a therapeutic target in AH decades ago.^51^ Since that time, preclinical and clinical studies have identified inflammatory pathways and mediators that contribute to liver damage and hepatocyte death. The cytokine TNFα is among the inflammatory mediators most strongly linked to AH. Patients with AH have higher levels of TNFα in their blood than healthy controls and TNFα is increased in the liver of animals chronically consuming ethanol versus control animals.^52–54^ At the cellular level, ethanol stabilizes TNFα mRNA in macrophages, leading to rapid increases in TNFα production and release from these cells.^48, 55, 56^ Macrophages also produce TNFα in response to MAMPs and DAMPs that are increased in CAC.^57^ Alcohol increases MAMP delivery to the liver by damaging the intestinal epithelium and allowing translocation across the gut mucosal barrier.^58^ Alcohol also increases DAMPs in the liver due to secretion from stressed cells and release from dead or dying cells.^15^ The high level of TNFα generated in these conditions activates programmed cell death in hepatocytes both *in vitro* and *in vivo*.^46, 59^ In pre-clinical models, TNF receptor 1 knockout or anti-TNFα antibody protect animals from liver damage.^54, 60^ However, clinical studies of anti-TNFα antibodies were halted early due to life-threatening infections in the AH patients.^61, 62^ This highlights the importance of targeting liver damaging “sterile inflammation” while leaving host defense intact in AH.

The first step in targeting liver damaging sterile inflammation is identifying appropriate therapeutic targets and biomarkers for those targets. Our findings suggest that HSPB1 is a component of sterile inflammation in AH and therefore warrants further investigation as a biomarker and/or therapeutic target in this disease. HSPB1 fits several criteria for these applications: 1) Differences in HSPB1 can be measured in the blood. 2) HSPB1 is significantly higher in AH than other forms of liver disease and increases with increasing disease severity. 3) HSPB1 is upstream of liver-damaging inflammation and so potentially indicates risk for inflammation before it occurs. 4) Blocking HSPB1 action blocks TNFα mediated inflammation *in vitro*.

HSPB1 has already been proposed as a biomarker for disease severity in cancers, most notably hepatocellular carcinoma (HCC).^27^ In HCC, HSPB1 is increased in lymphomonocytes and tumor tissue, especially in cases associated with a history of alcohol use.^28, 47^ High levels of liver HSPB1 have also been associated with increased immune cell infiltration into tumor tissue and worse prognosis in HCC.^27, 28^ Alcohol-associated liver diseases have also been linked to naturally occurring anti-HSPB1 antibodies.^25^ These antibodies could be important both diagnostically and therapeutically. Concern has been raised that anti-HSPB1 antibodies could interfere with measurement of HSPB1 in standard, ELISA-based assays.^31^ Additionally, naturally occurring anti-HSPB1 antibodies could influence HSPB1 functions and/or interfere with antibody-based therapeutics directed at HSPB1. Here, we utilized a commercially available ELISA, and results supported that this assay is able to measure the form and amount of HSPB1 released from ethanol stressed cells. We did not measure anti-HSPB1 antibodies in our patient population due to limited sample volume and this limitation also precluded any investigation of effects of naturally occurring anti-HSPB1 antibody on extracellular HSPB1 functions. These types of studies will need to be performed in the future if HSPB1 is developed as a therapeutic target for AH.

Development of HSPB1 as a biomarker for AH will also require prospective studies that measure HSPB1 concentration throughout the course of disease. High levels of HSPB1 may predict which patients are at highest risk for disease exacerbation and most in need of intense monitoring or early liver transplantation. Conversely, low HSPB1 may be useful to stratify patients into a low-risk group, since HSPB1 was no different in our patients consuming large amounts of alcohol without liver disease and healthy controls. Multiple timepoint sampling would reveal whether a single cut-off value of HSPB1 or changing levels of circulating HSPB1 would be most useful clinically.

Aside from its potential to serve as a biomarker in ALD, HSPB1 also shows promise as a therapeutic target in the inflammation of AH. Our data show that HSPB1 is upstream of TNFα mediated inflammation. Extracellular HSPB1 has also been reported to activate a second macrophage inflammatory mediator implicated in AH, IL-1β. Therefore, targeting HSPB1 has the potential to block activation of multiple inflammatory pathways and broadly protect hepatocytes from inflammation activated cell death programs. Our work specifically demonstrates the potential efficacy of an anti-HSPB1 antibody to interrupt inflammation in AH. We would expect this type of agent to be particularly effective at blocking the DAMP function of HSPB1, while leaving intracellular, cytoprotective functions of HSPB1 intact. It would also preserve TNFα activation by microbes and allow TNFα mediated inflammation in host defense mounted by macrophages.

Together, our findings directly link HSPB1 to inflammation in AH and provide a potential explanation for disease exacerbation. We propose that HSPB1 is highly upregulated in hepatocytes in response to ethanol metabolism and released into the extracellular environment where it activates TNFα in macrophages. When ethanol exposure is limited in amount or duration, we presume that HSPB1 is both cytoprotective in hepatocytes and promotes tissue repair through activation of low levels of TNFα at the site of damage. Conversely, when ethanol exposure occurs at high levels or is sustained for long periods of time, the high levels of TNFα could overwhelm HSPB1 cryoprotection, leading to hepatocyte death. Additionally, the dying ethanol stressed hepatocytes would be expected to release high levels of HSPB1, which would amplify TNFα production and causes even more hepatocyte death leading to a feed-forward or vicious cycle. Our data support this model in a number of ways. However, several key questions do remain unanswered. First, does extracellular HSPB1 act on cells other than macrophages during CAC? Hepatic stellate cells and other infiltrating innate and adaptive immune cells could also be influenced by HSPB1 to contribute to disease. Second, does anti-HSPB1antibody inhibit inflammation in the complex context of the intact liver during CAC? To answer this question, we need to test our anti-HSPB1 antibody in an animal model that mimics more severe, inflammatory ALD. This experiment would also reveal any negative effects of anti-HSPB1 antibody on hepatocyte health in this context. Lastly, which inflammatory pathways does extracellular HSPB1 activate in the liver during CAC? We focused on TNFα in this study due to its strong link to AH, but HSPB1 likely also activates IL-1β and potentially other pro-inflammatory cytokines in CAC. Answering these questions could lead to novel diagnostic and therapeutic approaches to ALD based on targeting HSPB1. This type of strategy could identify patients at risk of severe inflammatory liver disease and short-circuit inflammation to protect the liver against life-threatening injury.

## MATERIALS AND METHODS

### Human subjects

Study protocols were approved by the Institutional Review Boards (IRB) at the Cleveland Clinic and MetroHealth Hospitals. All procedures were performed according to the IRB guidelines and all subjects provided written informed consent.

#### Human serum samples

Enrolled patients had confirmed diagnosis of ALD by clinicians based on medical history, physical examination, and laboratory results, according to the guidelines of the American College of Gastroenterology (https://gi.org/clinical-guidelines/). Healthy controls (HC) were recruited from the Clinical Research Unit at the Cleveland Clinic. Serum was collected in gold top tubes and allowed to clot for 30 minutes at room temperature. Samples were then centrifuged for 10 minutes at 4°C. Aliquots were frozen at −80°C until use. Descriptive demographic and clinical data from these patients are provided in Tables 1-3.

#### Human liver samples

Healthy control and alcohol-associated hepatitis liver samples were provided by the NIAAA R24 Clinical Resource for Alcohol-associated Hepatitis Investigations at Johns Hopkins University. The resource provided liver from 5 patients with severe AH and 5 healthy donors that was obtained during liver transplantation. Tissues were snap frozen in liquid nitrogen and stored at −80°C prior to use. Descriptive demographic and clinical data from these patients are provided in Table 4.

#### Human liver RNA-seq data and analysis

RNA transcipts were obtained from the InTEAM Consortium – Alcoholic Hepatitis dbGaP Study, Accession phs001807.v1.p1. Transcripts per million (TPMs) for visualization were calculated as follows. Raw counts were normalized by dividing the counts by the length of each gene in kilobases to obtain reads per kilobases (RPK). The total RPK values in a sample were summed and then divided by 1,000,000 to give the “per million” scaling factor. Log2 fold changes were calculated by running raw counts through the DESeq2 pipline in R (v 4.2.2).

### Animal studies

All animal protocols were approved by the Cleveland Clinic Institutional Animal Care and Use Committee. Mice used in this study were C57BL/6 mice (Strain 000664) purchased from Jackson Laboratories and maintained under specific pathogen free (SPF) conditions. Mice were allowed to acclimate to the Cleveland Clinic animal facility for 5 days prior to the start of studies. All animal experiments were performed two independent times using female mice between 6 and 12 weeks of age. Female mice were chosen because of published data demonstrating that ALD is more severe in female mice and humans. Mice were housed under a 12-hour light/dark cycle in shoe-box cages (2 animals/cage) with microisolator lids and fed standard laboratory chow. Standard microisolator handling procedures were used throughout the study.

#### Lieber-DeCarli Diet model of chronic alcohol consumption

Mice were randomized into ethanol-fed (EtOH) and pair-fed (Pair/Control) groups and then adapted to the base liquid diet (Dyets, Bethlehem, PA; 710260) for 2 days. The EtOH group was then allowed free access to the base diet that contained increasing concentrations of EtOH over time: 1% and 2% (vol/vol) each for 2 days, then 4 and 5% EtOH each for 7 days, and finally 6% EtOH for another 7 days. The 6% (vol/vol) diet provided EOH as 32% of total calories in the diet. Control mice were pair-fed diets which isocalorically substituted maltodextrin for EtOH over the entire feeding period. At the end of the study, mice were anesthetized, blood samples were obtained from the posterior vena cava using nonheparinized syringes, livers were excised, and the mice were euthanized by exsanguination.

#### Mouse liver protein isolation

Snap frozen liver sections were placed into microcentrifuge tubes with 400 μL of tissue lysis buffer (Cell Signaling, 9803S) supplemented with protease inhibitor (Roche, 11836170001) and placed on ice. Livers were homogenized using a Tissue-Tearor. Protein concentration was determined by BCA (Pierce, 23228).

### Cell culture studies

All cell lines were obtained directly from American Type Culture Collection (ATCC) or from the Cleveland Clinic Cell Culture Core which obtained them directly from ATCC. Cell phenotypes were routinely monitored, and cells were periodically tested for mycoplasma contamination.

#### HSPB1 recombinant protein

HEK293 cells were transfected with an endotoxin-free prep (Takara Bio, 740426.50) of HSPB1-FLAG DNA (Genecopoeia, EX-I0586-M39) using TransIT-293 (Mirus Bio, MIR2705) according to the manufacturer’s protocol. Cells were transfected for 72 hours, then lysate was collected in TBS + 1 mM DTT supplemented with 1x Protease Inhibitor Cocktail (Roche, 11836170001). Lysates were sonicated on ice (1 min total time on; 10 seconds on/10 seconds off; Amp= 30%) and centrifuged at 15,000 x G for 20 min at 4 °C. The supernatant was then dialyzed overnight into TBS + 0.15 M NaCl, pH 7. The dialyzed supernatant was then directly applied onto 1mL bed of pre-equilibrated ANTI-FLAG® M2 Affinity Gel resin (Sigma, A2220). After allowing the protein to bind by gravity flow, the column was washed with 15 column volumes of TBS. HSPB1-FLAG was eluted by pouring five 1 bed volumes worth of 0.1M Glycine-HCl, pH 3.5. Five elution fractions were collected in 5 sequential 1.5 mL microcentrifuge tubes, each containing 50 µl of 0.1M Tris pH 8.0 to neutralize acid. Elution fractions were pooled together and spun down in 10 kDa spin concentrator to increase concentration, then buffer exchanged into HEPES storage buffer. Protein was then stored at 4°C.

#### Ethanol stress in hepatocytes

HepG2 hepatocytes were seeded into a 6-well tissue culture plate at 250,000 cells per well. The following day, the media was removed from the wells and the cells were treated with either 500 μL of 2.5 mM (1.5%) ethanol in serum-free EMEM or 500 μL serum-free EMEM for 1 hour at 37°C and 5% CO_2_. After ethanol stress, the supernatant was removed from the cells and frozen at −80°C until use. The cells were then scraped off the plate with a disposable cell scraper into 300 μL of PBS, pipetted up and down to homogenize lysate, and then frozen at −80°C until use.

#### Hepatocytes stressed with ethanol in the presence of 4-methylpyrazole (4-MP)

HepG2 or AML12 hepatocytes were seeded into a 6-well tissue culture plate at 250,000 cells per well. The following day, the media was removed from the wells, and the cells were treated with 500 μL of 2.5 mM (1.5%) ethanol in serum-free EMEM, 5 mM 4MP in 500 μL of serum-free EMEM, or 500 μL serum-free EMEM for 1 hour at 37°C and 5% CO_2_. After incubation, the supernatant was removed from the cells and frozen at −80°C until use. The cells were then scraped off the plate with a disposable cell scraper into 300 μL of PBS, pipetted up and down to homogenize lysate, and then frozen at −80°C until use.

#### Ethanol stress in macrophages treated with recombinant human HSPB1

RAW264.7 macrophages were seeded into a 6-well tissue culture plate at 250,000 cells per well. The following day, the media was removed from the wells and the cells were treated with 500 μL of 2.5 mM (1.5%) ethanol in serum-free DMEM, 3 μM HSPB1 in 500 μL of serum-free DMEM, 3 μM HSPB1, and 2.5 mM ethanol in serum-free DMEM, or 500 μL of DMEM for 1 hour or 24 hours at 37°C and 5% CO_2_.

#### Macrophages exposed to media conditioned by ethanol stressed hepatocytes and treated with anti-HSPB1 antibody

Two T75 tissue culture flasks containing HepG2 hepatocytes (~70-80% confluent) were treated with 10 mL of serum-free DMEM or 10 mL of serum-free DMEM containing 2.5 mM (1.5%) ethanol for 24 hours at 37°C and 5% CO_2_. The conditioned media was then removed from the culture flasks and split in half. Half of the media was used as a control and the other half was incubated with 2.5 mg/mL anti-HSPB1 antibody (Custom antibody produced by Biomatik). Both control and antibody treated media samples were rotated overnight at 4°C. RAW264.7 macrophages were plated the day prior to conditioned media collection at 250,000 cells per well in a 6 well tissue culture dish in DMEM supplemented with 10% FBS. After preparation of the treated and control conditioned media, the complete media was removed from the RAW264.7 cells. The RAW264.7 cells were then treated with 500 μL of serum-free DMEM (negative experimental control), serum-free DMEM with 2.5mM ethanol (positive experimental control), non-ethanol treated HepG2 conditioned media, non-ethanol treated HepG2 conditioned media with anti-HSPB1 antibody, ethanol treated HepG2 conditioned media, or ethanol treated HepG2 conditioned media with anti-HSPB1 antibody for 1 hour at 37°C and 5% CO_2_.

### HSPB1 assays

#### HSPB1 and phosphorylated-HSPB1 in human serum by ELISA

For measurement of HSPB1, serum samples were diluted 1:4 in reagent diluent and the ELISA completed per the manufacture’s protocol (R&D, DY1580). For measurement of phosphorylated-HSPB1 (S78/S82), serum samples were diluted 1:2 in reagent diluent and the ELISA completed per the manufacture’s protocol (R&D, DYC2314-2).

#### HSPB1 in murine liver by ELISA

100 μL of lysate (normalized by protein concentration) was loaded in triplicate onto a high-binding polystyrene plate (Costar, 07-200-39) and incubated overnight at 4°C. The following day, lysate was decanted, and the plate washed once with 200 μL per well of PBS with 0.01% Tween-20. The plate was blocked using 1% BSA in PBS (200 μL per well) for 1.5 hours at room temperature. After blocking, 50 μL of 0.63 mg/mL anti-HSPB1 antibody (Biomatik) was added to the plate and incubated for 2 hours at room temperature. After primary antibody incubation, the plate was washed 3 times with 200 μL of PBS with 0.01% Tween-20 per well. Then 50 μL of secondary antibody conjugated with horseradish peroxidase (Jackson Immunoresearch, 711-035-152) was added to the plate at a final concentration of 0.08 mg/mL for 1 hour. The plate was then again washed 3 times with PBS with 0.01% Tween-20 and 50 μL of pre-warmed TMB was added to the plate. The detection reagent was developed for 5 minutes at room temperature and then the reaction was quenched with 50 μL of 0.5M sulfuric acid. The plate was then read at 450_nm_ on a spectrophotometer (Synergy).

#### qRT-PCR for HSPB1 in murine liver

Total RNA was isolated from snap frozen liver samples by Trizol:chloroform extraction as per the manufacturer’s protocol (Invitrogen, 15596026), then precipitated using glycoblue, 3M sodium acetate and isopropanol. After washing twice with 80% ethanol, the RNA pellet was resuspended in nuclease free water and further purified using an RNA Clean and Concentrate column as per manufacturer protocol (Zymo Research, R1013). 1 mg of total RNA per sample was reverse transcribed (SuperScript IV Reverse Transcriptase, oligo dT primers, ThermoFisher, 18091050), and quantitative PCR performed on a QuantStudio 6 Flex Real-Time PCR System (Applied Biosystems) using TaqMan Fast Advanced Master Mix (Applied Biosystems, 4444557) and TaqMan probes for *Hspb1* (ThermoFisher, MM00834384_G1) and *Gapdh* (ThermoFisher, MM99999915_G1). *Hspb1* transcript levels were normalized to *Gapdh* and fold changes were calculated using the delta-delta CT method.

#### qRT-PCR for HSPB1 in HepG2 cells

At the end of experiments described above, media was removed from the cells, and cells were gently rinsed once with 500 μL of DPBS (Dubucco’s phosphate buffered saline, Genesee, 25-508). DPBS was removed and 200 μL of RNAlater (Invitrogen, AM7020) was added to each well. Cells were scraped off and collected into a microcentrifuge tube. RNA was isolated using PureLink RNA mini kit (Ambion, 12-183-018A) per the manufacturer’s instructions. 1 mg of total RNA per sample was reverse transcribed (Applied Biosystems, 43-688-14) and quantitative PCR performed on a CFX96 Touch Real-Time PCR system (Bio-Rad). Primer sequences: *Hspb1* Forward - 5’AAGCTAGCCACGCAGTCCAA3’; *Hspb1* Reverse - 5’CGACTCGAAGGTGACTGGGA3’; *Gapdh* Forward - 5’TGGTATCGTGGAAGGACTCATGAC3’; *Gapdh* Reverse - 5’ATGCCAGTGAGCTTCCCGTTCAGC3’. Transcript levels were normalized to *Gapdh* and fold changes were calculated using the delta-delta CT method.

#### Immunostaining for HSPB1 in murine liver

Formalin-fixed paraffin embedded liver was sliced into 5 μm sections and placed on glass slides. The slides were heated to 60°C on a heat block for 30 minutes and then submerged 3 times for 5 minutes in xylene. The slides were then rehydrated by submerging them 30 times in 95% ethanol followed by 15 submersions in 70% ethanol, and finally, submersion for 1 minute in deionized water. Slides were washed 2 times in TBS with 0.01% Tween-20 for 5 minutes and then underwent antigen retrieval by steaming submerged in sodium citrate for 20 minutes followed by a 1-hour cooling period. After steaming, a hydrophobic border was drawn around the tissue with a PAP pen and slides were blocked with Superior blocking buffer (G-Biosciences, 786-660) for 1 hour at room temperature. Anti-HSPB1 antibody (Biomatik) was added to the slides at a final concentration of 1.26 mg/mL and left to incubate overnight at 4°C in a humid chamber. The following day, slides were washed for 15 minutes total in TBS with 0.01% Tween-20. Anti-rabbit Alexa Fluor 647 conjugated secondary antibody was added to the slides at a final concentration of 0.9 mg/mL and incubated for 30 minutes at room temperature protected from light (protection from light continued from this point forward). Slides were washed again for 15 minutes total in TBS with 0.01% Tween-20 and then counterstained with 10 mg/mL bisBenzimide H (Sigma-Aldrich, 33258) dissolved in TBS for 20 minutes at room temperature. Finally, slides were washed for 5 minutes with deionized water and cover slipped. All immunofluorescent sections were imaged using either a widefield fluorescent microscope (Keyence BZ-800) or an inverted confocal microscope (40x/1.25 oil objective on a Leica TCS-SP8-AOBS inverted confocal microscope (Leica Microsystems, GmbH, Wetzlar, Germany)).

#### HSPB1 in hepatocytes by ELISA

Cell culture media or cell lysates were thawed on ice and then normalized to protein content by BCA assay (Pierce, 23228). HSPB1 concentration was measured in human cells (HepG2) with the R&D Human HSP27 DuoSet ELISA kit (DY1580) according to the manufacturer’s protocol. HSPB1 concentration was measured in murine cells (AML12) using the same protocol described for the murine liver samples.

#### Immunoblotting for HSPB1

Cell or liver lysates were boiled in Laemmli buffer containing 10% b-mercaptoethanol for 10 minutes. SDS-PAGE Gel electrophoresis was performed using 12% NuPage Bis-Tris gels with MOPS buffers. Proteins were transferred to PVDF membranes and blocked with LiCOR Intercept PBS blocking buffer (LiCor, 927-70003) for 1 hour. Membranes were then incubated in blocking buffer containing 0.63 mg/mL anti-HSPB1 antibody (Biomatik) and 0.282 mg/mL anti-GAPDH antibody (Cell Signaling, D4C6R) overnight at 4°C with agitation. The membranes were washed 3 times with PBS with 0.01% Tween-20 for 5 minutes each wash and then incubated with 0.2 mg/mL 680RD (LI-COR, 926-68023) anti-rabbit or 0.2 mg/mL 800RD (LI-COR, 926-32212) anti-mouse secondary antibody in blocking buffer for one hour. Membranes were then again washed 3 times with PBS with 0.01% Tween-20 for 5 minutes and fluorescent signal was detected using an LI-COR Odyssey CLX machine and LI-COR Acquisition software. Densitometry was performed using ImageJ.

### Other assays

#### Liver Triglycerides

Measured pieces of liver (80-85mg) were digested in 3 M KOH at 70°C for 1 hour then left overnight at room temperature. The next day the total volume was brought to 500 μL with 2M Tris-HCl pH 7.5 and then diluted 1:5 with 2M Tris-HCl pH 7.5. 10 μL of diluted sample was added to of 1 mL of pre-warmed GPO triglyceride reagent (Pointe Scientific Inc., T7532-500). After a 5-minute incubation, samples were read on a spectrophotometer at 500 nm.

#### Aspartate aminotransferase (AST) and Alanine aminotransferase (ALT)

For both assays, murine serum was thawed on ice. For AST, 5 μL serum was added to 100 μL of AST reagent (Sekisui Diagnostics, 319-30) and read on a spectrophotometer at 340nm every 30 seconds for 20 minutes. For ALT, 10 μL serum was added to 90 μL of ALT reagent (Sekisui Diagnostics, 318-30) and read on a spectrophotometer at 340nm every 30 seconds for 30 minutes.

#### TNFα ELISA in cell culture supernatants

At the end of experiments described above, cell media was collected and TNFα concentrations measured using the Mouse TNF-alpha Quantikine ELISA Kit (R&D, #MTA00B) according to manufacturer’s protocol and recommendations.

### Statistics

Values shown in all figures represent the means ± SD, except figures 2C, D, and E which represent the means ± SEM. Pairwise differences were evaluated using parametric or non-parametric two-tailed t-tests as appropriate. Differences among multiple groups was detected using one-way ANOVAs with post-hoc analysis. Statistical analysis was performed using GraphPad Prism (v.10.2.2), except analysis in figures 2C-E, which was performed using R (v 4.2.2). A *P* value of less than 0.05 was considered significant.

## Data Availability

All data produced in the present study are available upon reasonable request to the authors.

## Conflict of Interest

The authors declare no conflicts of interest.

## Financial Support

NIDDK 1K08DK114713 (JSM), NIAAA Northeast Ohio Alcohol Research Center Pilot and Feasibility Award (JSM), NIAAA Northeast Ohio Alcohol Research Center 5P50AA024333 (LEN), NIAAA Johns Hopkins Clinical Resource for Alcoholic Hepatitis Investigation 5R24AA025017.

## Author Contributions

Conceptualization: JSM

Methodology: JSM, AMCO, MB, DR, AMS, LEN

Investigation: AMCO, MB, AB, MM, DC, EH, VP, CF, RV, SD, JD, DS, NW

Funding acquisition: JSM

Project administration: JSM

Supervision: JSM, AMCO, DR, AMS, LEN

Writing – original draft: JSM

Writing – review & editing: JSM, AMCO, MB, AMS, LEN

## Acknowledgements

We thank Connor Hansen for assistance with manuscript preparation. The NIAAA funded Northeast Ohio Alcohol Research Center Clinical and Pre-clinical Cores were instrumental to this work. We also acknowledge the assistance of the Cleveland Clinic Light Microscopy Core in this study.

## Data and materials availability

All data and code used in the analysis are available upon request. Materials used in this study are available upon request and with completion of the appropriate material transfer agreements.

## Notes

### Competing Interest Statement

The authors have declared no competing interest.

### Funding Statement

This study was funded by NIDDK 1K08DK114713 (JSM), NIAAA Northeast Ohio Alcohol Research Center Pilot and Feasibility Award (JSM), NIAAA Northeast Ohio Alcohol Research Center 5P50AA024333 (LEN), NIAAA Johns Hopkins Clinical Resource for Alcoholic Hepatitis Investigation 5R24AA025017.

### Author Declarations

Study protocols were approved by the Institutional Review Boards (IRB) at the Cleveland Clinic and MetroHealth Hospitals. The IRB of Cleveland Clinic and MetroHealth Hospitals gave ethical approval for this work. All procedures were performed according to the IRB guidelines and all subjects provided written informed consent.

